# Complementary and Alternative Medicine Therapies in relation to Self-Reported Better Sleep across Racial and Ethnic Groups in the United States

**DOI:** 10.64898/2026.09.04.26362273

**Authors:** Rupsha Singh, Symielle A. Gaston, Christopher Payne, Dayna T. Neo, Suzanne M. Bertisch, Wayne B. Jonas, Chandra L. Jackson

## Abstract

**Objectives:** Complementary and Alternative Medicine (CAM) approaches vary across race and ethnicity, reflecting different cultural practices. However, its relationship with sleep is understudied. We investigated cross-sectional CAM-sleep associations across racial and ethnic groups.

**Methods:** In the nationally-representative 2012 National Health Interview Survey, participants identified their most important CAM therapies and whether the therapy yielded ‘better sleep’ (yes/no). We used Poisson regression with robust variance to estimate adjusted prevalence ratios (aPRs) and 95% confidence intervals (CIs) and examined effect measure modification using Wald tests of interaction terms.

**Results:** Among 9,308 adults (mean age±SE = 46.4±0.3 years), 58.2% were women. CAM therapy prevalence ranged from 0.1% (craniosacral therapy) to 28.9% (herbal supplements). The proportion perceiving ‘better sleep’ was 41.3%. When deemed most important for health, massage; meditation; movement or exercise techniques; and yoga, Tai Chi, and Qigong were associated with higher prevalence of perceiving ‘better sleep’ similarly across racial and ethnic groups. Chiropractic or osteopathic manipulation was associated with perceiving better sleep only among non-Hispanic (NH)-White adults (aPR=1.14, 95% CI:1.05-1.24). The highest magnitude of associations between herbal supplement use and lower prevalence of ‘better sleep’ was among NH-White adults (aPR=0.29, 95% CI:0.25-0.34 vs. aPR range:0.37 (NH-Asian) to 0.50 (NH-Black)). Limited by a small sample size, no associations were observed among NH-American Indian/Alaska Native participants.

**Conclusion:** Many CAM therapies were associated with perceiving better sleep across racial and ethnic groups; however, effectiveness varied by therapy and racial and ethnic groups, yielding mixed evidence of their potential role in sleep health promotion.

## INTRODUCTION

Insufficient sleep and sleep disorders are highly prevalent, affecting one-third of the US population and approximately 50-70 million Americans, respectively (1, 2). Poor sleep health is associated with adverse mental health, lower life satisfaction, poor cardiometabolic health (e.g., type 2 diabetes), and premature mortality (3-6). Certain racial and ethnic groups generally demonstrate poorer sleep than other groups. For instance, Non-Hispanic (NH)-Black/African American adults –compared to NH-White counterparts– experience a higher burden of short sleep duration and lower subjective sleep quality while Hispanic/Latino and Asian adults have a higher prevalence of short sleep duration (7). Sleep disparities likely contribute to other well-documented mental and physical health disparities (8). While behavioral and pharmacological interventions can improve sleep, their use may be limited by affordability, access to sleep specialists, pharmacological side effects, and patient preferences (9, 10). Additionally, there is little evidence about long-term sleep medication benefits (11). Accordingly, interest has grown in alternative approaches such as Complementary and Alternative Medicine.

Complementary and Alternative Medicine (CAM) can supplement (‘complementary’) or substitute (‘alternative’) conventional treatment (12-14). CAM includes a variety of products and practices rooted in holistic healing and natural remedies to promote relaxation and improve well-being. CAM therapies include mind-body therapies (e.g., meditation, guided imagery), energy-based therapies (e.g., reiki, healing touch), manipulative and body-based practices (e.g., reflexology and massage), biologically-based practices (e.g., dietary supplements and botanicals), and whole medical systems (e.g., Ayurvedic and traditional Chinese medicine) (15). Increasingly, whole-person approaches to sleep health recognize that sleep is influenced not only by biological and behavioral factors, but also by psychological, social, cultural, and spiritual dimensions of health, many of which overlap with CAM (16). Recently, adults have shown increased interest in CAM to improve sleep onset, maintain sleep continuity, and reduce physical discomfort that disrupts rest (17-19). Consistent with this interest, studies have demonstrated that mindfulness-based interventions, including mindfulness meditation, yoga, Tai Chi, and Qigong, can improve subjective sleep quality, although these have not consistently outperformed conventional treatments such as cognitive behavioral therapy for insomnia (17, 18). Nonetheless, these therapies may improve sleep health by promoting relaxation through reducing anxiety and stress, enhancing coping resources, and improving emotional regulation, mechanisms consistent with whole-person integrative care frameworks that recognize the interconnected spiritual, biological, psychological, behavioral, and social dimensions of health and well-being (19-25). Additionally, manipulative and body-based therapies are hypothesized to reduce pain or musculoskeletal discomfort that disrupt sleep (26). Together, these diverse CAM therapies offer holistic approaches that address both physical and psychological factors affecting sleep, underscoring their potential role in enhancing overall sleep health.

Evidence suggests that patterns of utilization and types of CAM therapies differ across racial and ethnic groups (27-30). In a prior study, the prevalence of CAM use was highest among NH-White adults followed by Hispanic and NH-Black adults (28). Regarding specific modalities, that study also showed that NH-White, NH-Black, and Hispanic adults most often reported herbal medicine and relaxation techniques (28), while in another study, Hispanic and Asian adults most often used herbal supplements, meditation, and traditional healing practices (29). Although utilization and type of CAM therapy varies by racial and ethnic groups, studies suggest that therapeutic benefits associated with CAM use do not mirror utilization patterns. For instance, a 2012 NHIS study of adults aged 50 years and older reported that although CAM was less common among NH-American Indian/Alaska Native (AI/AN), NH-Asian, NH-Black, and Hispanic adults, individuals in these groups who used CAM reported more health and wellness benefits compared to NH-White individuals (31). One hypothesis is that differential treatment across racial and ethnic groups within conventional healthcare systems may lead some individuals to seek CAM as a more patient-centered (or directed) and culturally-responsive approach (32). As such, variations in the prevalence and types of CAM use may lead to differences in sleep-related benefits across racial and ethnic groups.

While prior studies have examined CAM use among individuals with sleep difficulties (33), as well as racial and ethnic differences in perceived sleep benefits of broadly using CAM (31), little is known about differences in perceived sleep-related benefits from specific CAM therapies across racial and ethnic groups. Of the few studies, one found that adults with insomnia symptoms were more likely to use CAM therapies (33), another study demonstrated perceived sleep health benefits with use of any (non-specific) CAM therapies (34), and another showed racial and ethnic differences in perceived sleep benefits from using any CAM (31). However, these studies did not examine sleep-related benefits across specific CAM modalities and racial and ethnic groups simultaneously. Thus, we investigated racial and ethnic differences in self-reported better sleep from use of individual CAM therapies. We hypothesized that CAM therapies are associated with better sleep overall and associations will be stronger among NH-AI/AN, NH-Asian, NH-Black, and Hispanic/Latino adults compared to NH-White adults.

## METHODS

### Data Source

Using the Integrated Public Use Microdata (IPUMS) (35), we analyzed cross-sectional data from the 2012 National Health Interview Survey (NHIS), a nationally representative survey of the non-institutionalized U.S. population that uses a multistage sampling design. In 2012, the most recent year with CAM and sleep data, sample adult interviews were completed by 34,525 participants (conditional response rate=79.7%) (36). All participants consented to the study, and approval for this study using de-identified, public data is waived by the National Institutes of Health Institutional Review Board.

### Study population

Participants who did not respond to the CAM supplement (n=931), who reported no CAM use (n=23,413), and who had missing information for self-reported better sleep and potential confounders (n=873) were excluded (Supplemental Figure 1). The final analytic sample included 9,308 participants aged 18 years and older who reported using at least one CAM therapy in the past 12 months. Compared with included participants, excluded participants were older, more likely to be women, identify as NH-Black or NH-White, have lower educational attainment, be unemployed or not in the labor force, and reside in the Northeast region of the U.S. (all *p*<0.05) but did not differ in general health status (Supplemental Table 1).

### Exposure Assessment: Complementary and Alternative Medicine Therapies

The 2012 NHIS Adult Complementary and Alternative Medicine Supplement asked participants questions about their use of CAM therapies. These therapies included acupuncture; ayurveda; biofeedback; chelation therapy; chiropractic or osteopathic manipulation; craniosacral therapy; energy healing therapy; herbal and non-vitamin supplements; homeopathy; hypnosis; massage; meditation, guided imagery, and progressive relaxation techniques; movement and exercise techniques; naturopathy; special diets; traditional healers; and yoga, tai chi, or qigong. Details about herbal and non-vitamin supplements, meditation and relaxation techniques, traditional healers, movement and exercise methods, and special diets are provided in the appendix (Appendix I-VI). For most CAM therapies, participants were asked whether they had ever used the therapy or were seen by a practitioner of the therapy, and if so, if it occurred within the past 12 months. Participants were also asked how often and why they used each therapy or visited a practitioner of the therapy. Costs associated with them were also assessed.

We focused on a series of questions that asked participants to identify three CAM therapies most important to their health. These therapies were ranked first, second, or third based on participant responses, except for ‘ayurveda’, ‘chelation therapy’, and ‘vitamins and minerals’ which were not included in the list. If reporting herbal and non-vitamin supplements, participants were asked to identify up to two supplements they used most frequently or considered most important for their health. For our analysis, two specific supplements (if reported) were not considered separately but rather combined generally as ‘herbal supplements’. Participants could only choose one CAM therapy as most important to their health. Therefore, for each CAM therapy, we considered whether participants identified that specific therapy as the most important for their health, as opposed to either not selecting it as most important or choosing another therapy as most important.

### Outcome Assessment: Perceived ‘better sleep’

Participants were asked about: 1) reasons for using each of the top three CAM therapies, 2) whether the therapies were used to treat a specific health condition, 3) which therapy was used most to treat those conditions, and 4) the physical and mental health outcomes associated with their use. Perceived better sleep from use of the CAM modality identified by participants as most important to their health was derived from a question asking participants if seeing a practitioner for [a particular modality] or using the modality led to any of these outcomes…Help you sleep better? (Yes, No). Responses of “Don’t know” and “Refused” were coded as missing.

### Potential confounders

Potential confounders, described in Table 1, were selected *a priori* based on prior literature and their potential associations with both CAM therapy use and sleep (37), and included age, sex, race and ethnicity, educational attainment, annual household income, employment status, marital status, region of residence, body mass index category, and general health status. We also examined other characteristics that were not identified as confounders, such as usual sleep duration, insomnia symptoms, smoking status, alcohol consumption, leisure-time physical activity, dyslipidemia, hypertension, prediabetes/diabetes, and serious psychological distress.

**Table 1.**
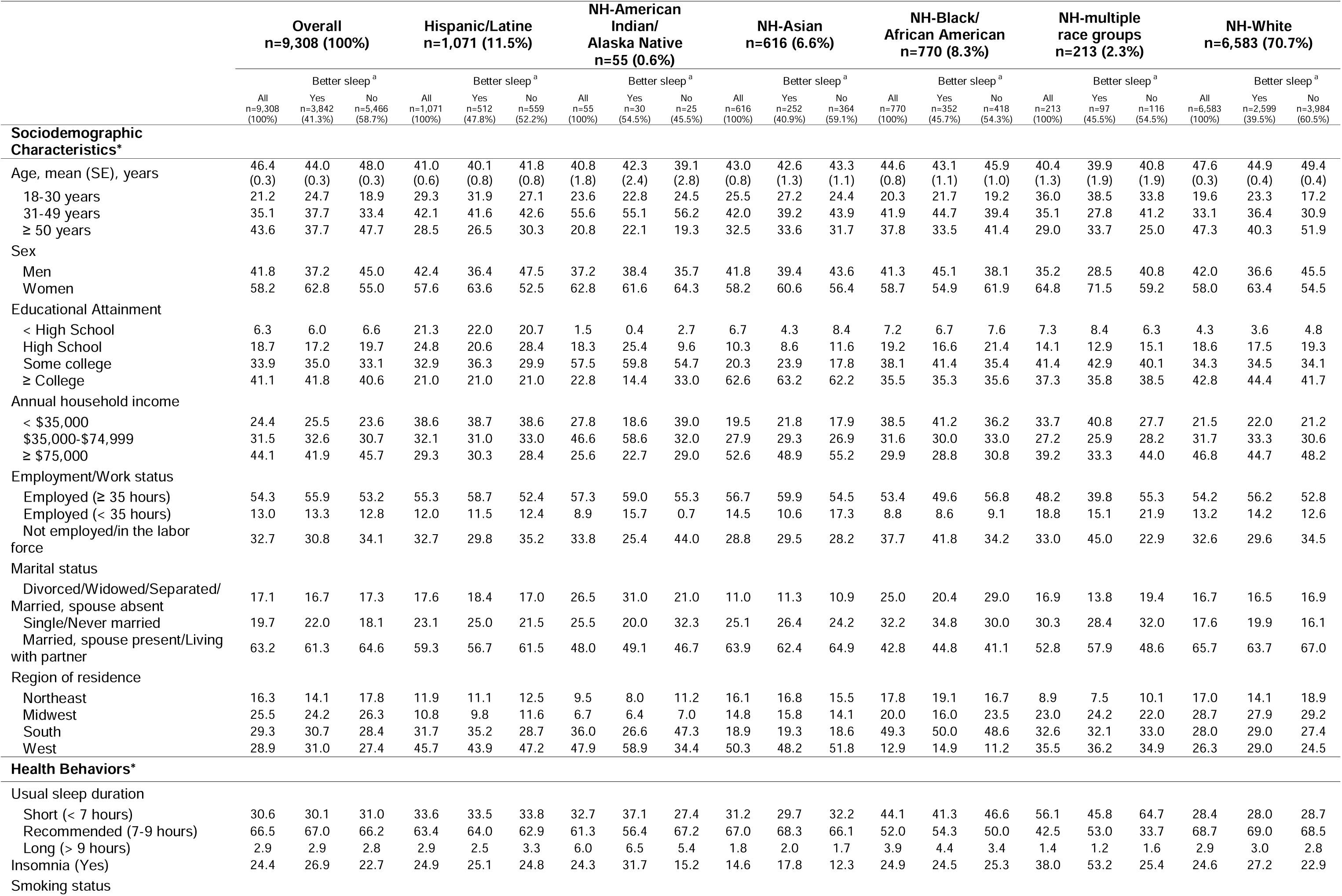
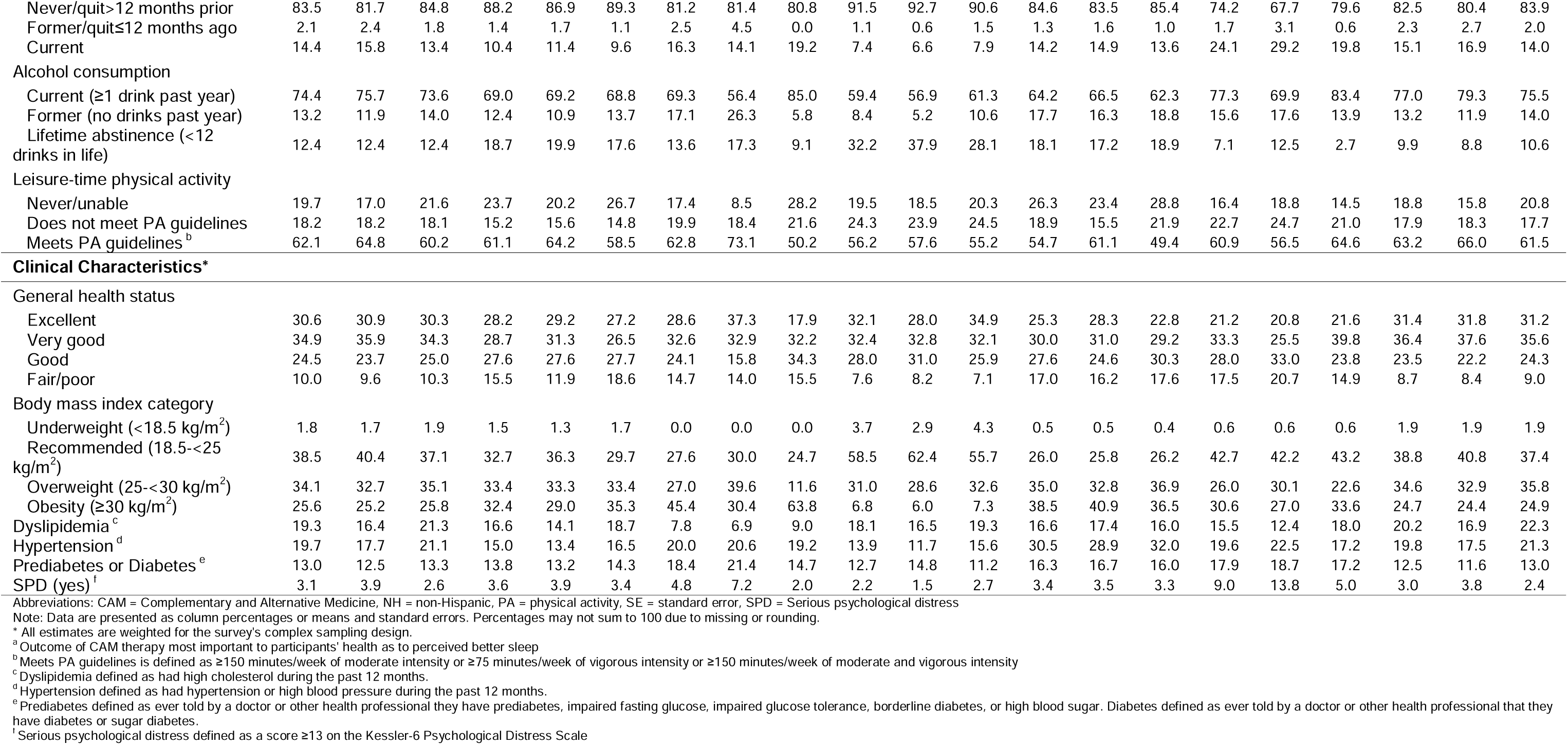
Study population characteristics of adults reporting the use of complementary and alternative medicine therapies most important to their health and perceived better sleep, overall and by race and ethnicity, National Health Interview Survey, 2012, (N=9,308)

### Potential effect modifier

Race and ethnicity (Hispanic/Latine, NH-American Indian/Alaska native, NH-Asian, NH-Black/African American, NH-multiple race groups, NH-White) was also included as a potential modifier.

### Statistical Analysis

Statistical analyses accounted for the National Health Interview Survey’s complex multistage sampling design by incorporating sampling weights, strata, and primary sampling units using survey procedures. The sampling weights account for participants’ probability of selection and include adjustments for survey nonresponse and post-stratification, allowing estimates to be nationally representative of the U.S. civilian noninstitutionalized adult population. Descriptive statistics were estimated overall, by ‘better sleep’, by race and ethnicity, and by ‘better sleep’ within racial and ethnic groups. Continuous variables are presented as survey-weighted means with standard errors (SEs), and categorical variables are summarized as survey-weighted percentages. Poisson regression with robust standard errors was used to estimate prevalence ratios and 95% confidence intervals for associations between individual CAM therapy use and better sleep adjusting for aforementioned confounders. Effect modification by race and ethnicity was assessed through multiplicative interaction terms in adjusted models within the overall population. Wald tests were used to evaluate interaction terms at a significance level of 0.05. All analyses were performed using Stata, Version 19.5 (Statacorp, College Station, Texas).

## RESULTS

### Study population characteristics

#### Overall

Among 9,308 participants who reported using at least one CAM therapy, the average age was 46.4 ± 0.3 years, and most participants identified as NH-White (76.4%), followed by Hispanic/Latine (9.7%), NH-Black/African American (6.2%), and NH-Asian (5.4%), NH-Multiple or other race groups (2.0%), and NH-American Indian/Alaska Native (0.5%). NH-Multiple or other race groups included all other racial groups in which sample sizes were too small to consider separately (Supplemental Table 1). Results for the NH-multiple or other race groups are shown but not interpreted due to higher within-group heterogeneity (combining multiple races) than the remaining groups. Most participants were women (58.2%), attained ≥some college (75.0%), reported an annual household income of <$75,000 (55.9%), were employed ≥ 35 hours/week (54.3%), were married or living with a partner (63.2%), reported habitually sleeping 7 to 9 hours (66.5%), and reported very good or excellent general health (65.5%) (Table 1).

#### By race and ethnicity

Nearly half (47.3%) of NH-White participants were aged ≥50 years, while participants of other racial and ethnic groups were predominantly 31-49 years old (Table1). While other sociodemographic and health characteristics were similar across groups, educational attainment and annual household income varied by race and ethnicity. Specifically, Hispanic/Latine, NH-Black/African American, and NH-AI/AN participants more frequently reported completing some college, whereas NH-Asian and NH-White participants more often reported completing ≥bachelor’s degree. Additionally, annual household incomes <$35,000 were more common among Hispanic/Latine and NH-Black/African American participants, while incomes ≥$75,000 were more common among NH-Asian and NH-White participants.

### Prevalence of CAM therapies prioritized as important for health

Table 2 presents the weighted prevalences of CAM therapies reported most important to health and perceived better sleep. The top five were herbal supplements (28.9%), chiropractic or osteopathic manipulation (25.7%), yoga, Tai Chi, or Qigong (13.4%), massage (13.1%), and meditation, guided imagery or progressive relaxation (6.8%). When stratified by race and ethnicity, the top five CAM therapies and patterns by sleep-related benefits were generally consistent with those observed in the overall population, except among NH-AI/AN participants, among whom 32.7% identified chiropractic and osteopathic manipulation and 30.8% identified traditional healers (vs. range among other groups: 0.1% (NH-White) to 6.2% (Hispanic/Latine)) as most important for health.

**Table 2.** Weighted Prevalence of the Use of Complementary and Alternative Medicine Therapies reported as most important to respondent’s health, Overall and by Race and Ethnicity and perceived better sleep, National Health Interview Survey, 2012, (N=9,308)

|  | Overall<br>n=9,308 (100%) |  |  | Hispanic/Latine<br>n=1,071 (11.5%) |  |  | NH-American<br>Indian/<br>Alaska Native<br>n=55 (0.6%) |  |  | NH-Asian<br>n=616 (6.6%) |  |  | NH-Black/<br>African American<br>n=770 (8.3%) |  |  | NH-multiple<br>race groups<br>n=213 (2.3%) |  |  | NH-White<br>n=6,583 (70.7%) |  |  |
| --- | --- | --- | --- | --- | --- | --- | --- | --- | --- | --- | --- | --- | --- | --- | --- | --- | --- | --- | --- | --- | --- |
|  | Better sleep<sup>a</sup> |  |  | Better sleep<sup>a</sup> |  |  | Better sleep<sup>a</sup> |  |  | Better sleep<sup>a</sup> |  |  | Better sleep<sup>a</sup> |  |  | Better sleep<sup>a</sup> |  |  | Better sleep<sup>a</sup> |  |  |
|  | All<br>n=9,308<br>(100%) | Yes<br>n=3,842<br>(41.3%) | No<br>n=5,466<br>(58.7%) | All<br>n=1,071<br>(100%) | Yes<br>n=512<br>(47.8%) | No<br>n=559<br>(52.2%) | All<br>n=55<br>(100%) | Yes<br>n=30<br>(54.5%) | No<br>n=25<br>(45.5%) | All<br>n=616<br>(100%) | Yes<br>n=252<br>(40.9%) | No<br>n=364<br>(59.1%) | All<br>n=770<br>(100%) | Yes<br>n=352<br>(45.7%) | No<br>n=418<br>(54.3%) | All<br>n=213<br>(100%) | Yes<br>n=97<br>(45.5%) | No<br>n=116<br>(54.5%) | All<br>n=6,583<br>(100%) | Yes<br>n=2,599<br>(39.5%) | No<br>n=3,984<br>(60.5%) |
| CAM Therapies*<sup>b</sup> |  |  |  |  |  |  |  |  |  |  |  |  |  |  |  |  |  |  |  |  |  |
| Acupuncture | 2.1 | 1.9 | 2.1 | 1.8 | 1.6 | 2.0 | 0.0 | 0.0 | 0.0 | 6.0 | 4.9 | 6.7 | 1.7 | 1.5 | 1.9 | 0.7 | 0.0 | 1.2 | 1.9 | 1.9 | 1.9 |
| Biofeedback | 0.5 | 0.5 | 0.5 | 0.2 | 0.2 | 0.2 | 0.0 | 0.0 | 0.0 | 0.0 | 0.0 | 0.0 | 0.8 | 1.3 | 0.4 | 1.2 | 0.0 | 2.3 | 0.6 | 0.5 | 0.6 |
| Chiropractic and<br>Osteopathic Manipulation | 25.7 | 26.2 | 25.3 | 20.5 | 18.9 | 21.9 | 32.7 | 40.1 | 23.7 | 10.9 | 6.3 | 14.1 | 14.9 | 12.4 | 17.0 | 18.3 | 10.6 | 24.7 | 28.4 | 30.5 | 27.0 |
| Craniosacral Therapy | 0.1 | 0.1 | 0.1 | 0.0 | 0.0 | 0.0 | 0.0 | 0.0 | 0.0 | 0.0 | 0.0 | 0.0 | 0.0 | 0.0 | 0.0 | 0.0 | 0.0 | 0.0 | 0.1 | 0.1 | 0.2 |
| Energy Healing Therapy | 0.4 | 0.6 | 0.3 | 0.6 | 1.1 | 0.1 | 0.0 | 0.0 | 0.0 | 0.2 | 0.0 | 0.3 | 0.3 | 0.3 | 0.3 | 1.5 | 3.3 | 0.0 | 0.4 | 0.5 | 0.3 |
| Herbal Supplements<sup>c</sup> | 28.9 | 11.7 | 40.7 | 27.2 | 14.8 | 38.0 | 12.1 | 2.0 | 24.4 | 29.1 | 13.0 | 40.5 | 33.9 | 20.5 | 45.3 | 28.2 | 21.6 | 33.7 | 28.8 | 10.1 | 40.9 |
| Homeopathy | 1.9 | 1.9 | 2.0 | 2.2 | 2.1 | 2.4 | 0.0 | 0.0 | 0.0 | 3.4 | 3.1 | 3.6 | 1.3 | 0.9 | 1.7 | 3.4 | 3.6 | 3.3 | 1.8 | 1.8 | 1.8 |
| Hypnosis | 0.5 | 0.4 | 0.5 | 0.4 | 0.0 | 0.7 | 0.0 | 0.0 | 0.0 | 0.0 | 0.0 | 0.0 | 0.0 | 0.0 | 0.0 | 0.0 | 0.0 | 0.0 | 0.6 | 0.6 | 0.6 |
| Massage | 13.1 | 17.5 | 10.2 | 14.5 | 17.7 | 11.7 | 5.7 | 5.5 | 5.9 | 14.4 | 20.9 | 9.9 | 12.9 | 17.6 | 8.9 | 6.9 | 9.9 | 4.5 | 13.1 | 17.5 | 10.3 |
| Meditation, guided<br>imagery, or progressive<br>relaxation | 6.8 | 12.2 | 3.1 | 6.3 | 10.5 | 2.8 | 11.8 | 17.3 | 5.1 | 6.2 | 10.7 | 3.1 | 8.7 | 16.4 | 2.2 | 10.4 | 14.5 | 6.9 | 6.6 | 12.0 | 3.2 |
| Movement or exercise<br>techniques | 1.5 | 2.5 | 0.8 | 1.5 | 2.9 | 0.3 | 0.0 | 0.0 | 0.0 | 1.4 | 1.7 | 1.3 | 2.4 | 3.6 | 1.4 | 1.0 | 2.2 | 0.0 | 1.4 | 2.5 | 0.8 |
| Naturopathy | 0.4 | 0.5 | 0.3 | 0.2 | 0.3 | 0.1 | 0.0 | 0.0 | 0.0 | <0.1 | 0.1 | 0.0 | 0.8 | 1.3 | 0.3 | 0.8 | 1.1 | 0.6 | 0.4 | 0.5 | 0.3 |
| Special diets | 3.7 | 4.0 | 3.5 | 4.2 | 4.9 | 3.6 | 1.9 | 3.5 | 0.0 | 5.3 | 5.1 | 5.5 | 5.6 | 5.0 | 6.0 | 6.8 | 4.6 | 8.7 | 3.3 | 3.7 | 3.1 |
| Traditional Healers | 0.9 | 0.8 | 0.9 | 6.2 | 5.1 | 7.1 | 30.8 | 22.5 | 40.9 | 0.4 | 0.0 | 0.7 | 0.2 | 0.1 | 0.4 | 1.6 | 0.6 | 2.5 | 0.1 | 0.2 | 0.1 |
| Yoga, Tai Chi, or Qigong | 13.4 | 19.0 | 9.6 | 14.2 | 20.0 | 9.2 | 5.0 | 9.0 | 0.0 | 22.5 | 34.2 | 14.3 | 16.4 | 19.1 | 14.1 | 19.2 | 28.1 | 11.7 | 12.4 | 17.5 | 9.0 |

Abbreviations: CAM = Complementary and Alternative Medicine, NH = non-Hispanic
Note: Data are presented as column percentages or means and standard errors. Percentages may not sum to 100 due to missing or rounding.
\* All estimates are weighted for the survey's complex sampling design.
<sup>a</sup> Outcome of CAM therapy reported as most important to respondent's health regarding perceived better sleep.
<sup>b</sup> CAM therapy reported as most important to respondent's health
<sup>c</sup> Includes combinations of herbal supplements

### Prevalence of perceived ‘better sleep’

Overall, the sample proportion perceiving ‘better sleep’ with use of any CAM modality deemed most important for health was 41.3% (Table 1). When stratified by race and ethnicity, the proportion was highest among NH-AI/AN (54.5%) and lowest among NH-White (39.5%) participants. Across all racial and ethnic groups, participants who reported sleep-related benefits was slightly younger or of similar age compared to those who did not report sleep-related benefits. The remaining sociodemographic, health behaviors, and sleep characteristics were generally similar between adults who did and did not report sleep-related benefits within each racial and ethnic group (except among NH-multiracial, which was not interpreted). The prevalence of perceived ‘better sleep’ for each specific CAM modality, overall and by race and ethnicity is shown in Supplemental Table 2.

### Unadjusted associations between CAM therapies and perceived ‘better sleep’

Overall, participants who reported sleep-related benefits were more likely to prioritize massage (17.5% vs. 10.2%), meditation, guided, imagery, or progressive relaxation (12.2% vs. 3.1%), and yoga, Tai Chi, or Qigong (19.0% vs. 9.6%) and less likely to prioritize herbal supplements (11.7% vs. 40.7%) as important for their health compared to participants who did not report sleep-related benefits (Table 2). Patterns were similar across racial and ethnic groups with few exceptions. In contrast to the remaining groups, NH-AI/AN participants who reported sleep-related benefits were slightly less likely to prioritize massage (5.5% vs. 5.9%) and notably less likely to prioritize herbal supplements (2.0% vs. 24.4%) compared with those who did not report ‘better sleep’.

### Adjusted associations between CAM therapy and perceived ‘better sleep’

#### Overall

Massage; meditation, guided imagery, or progressive relaxation; movement or exercise techniques; and yoga, Tai Chi, and Qigong were associated with a higher likelihood of perceiving ‘better sleep’ similarly across racial and ethnic groups (all p-interaction>0.05; Table 3). Prevalence ratios, adjusted for sociodemographic and clinical characteristics, ranged from 1.39 (massage: aPR=1.39 [95% CI:1.29-1.50]) to 1.82 (meditation, guided imagery, or progressive relaxation: aPR=1.82 [95% CI:1.69-1.97]).

**Table 3.** Prevalence ratios for associations between perceived better sleep and Complementary and Alternative Medicine Therapies reported as most important to respondent’s health, Overall and stratified by race and ethnicity, National Health Interview Survey, 2012, (N=9,308)

|  | Overall<br>n=9,308 (100%) |  | Hispanic/Latine<br>n=1,071 (11.5%) |  | NH-American<br>Indian/<br>Alaska Native<br>n=55 (0.6%) |  | NH-Asian<br>n=616 (6.6%) |  | NH-Black/<br>African American<br>n=770 (8.3%) |  | NH-multiple<br>race groups<br>n=213 (2.3%) |  | NH-White<br>n=6,583 (70.7%) |  |
| --- | --- | --- | --- | --- | --- | --- | --- | --- | --- | --- | --- | --- | --- | --- |
|  | Unadjusted | Adjusted <sup>a</sup> | Unadjusted | Adjusted <sup>b</sup> | Unadjusted | Adjusted <sup>b</sup> | Unadjusted | Adjusted <sup>b</sup> | Unadjusted | Adjusted <sup>b</sup> | Unadjusted | Adjusted <sup>b</sup> | Unadjusted | Adjusted <sup>b</sup> |
| Prevalence Ratio (95% Confidence Interval) for Associations with CAM therapies |  |  |  |  |  |  |  |  |  |  |  |  |  |  |
| CAM Therapies* <sup>b</sup> |  |  |  |  |  |  |  |  |  |  |  |  |  |  |
| Acupuncture | 0.95<br>(0.76-1.19) | 0.97<br>(0.78-1.21) | 0.89<br>(0.50-1.57) | 0.94<br>(0.52-1.70) | NE | NE | 0.82<br>(0.46-1.45) | 0.80<br>(0.45-1.39) | 0.85<br>(0.36-2.01) | 0.89<br>(0.41-1.96) | NE | NE | 1.01<br>(0.77-1.32) | 1.01<br>(0.77-1.32) |
| Biofeedback | 0.97<br>(0.63-1.50) | 1.03<br>(0.65-1.62) | 1.12<br>(0.34-3.66) | 1.18<br>(0.32-4.37) | NE | NE | NE | NE | 1.57<br>(0.83-2.98) | 1.65<br>(0.87-3.14) | NE | NE | 0.95<br>(0.57-1.59) | 1.02<br>(0.59-1.75) |
| Chiropractic or Osteopathic<br>Manipulation <sup>d</sup> | 1.03<br>(0.95-1.11) | 1.07<br>(0.99-1.16) | 0.90<br>(0.73-1.11) | 0.95<br>(0.78-1.16) | 1.38<br>(0.73-2.60) | 1.28<br>(0.69-2.38) | **0.55**<br>**(0.34-0.89)** | **0.52**<br>**(0.31-0.87)** | 0.81<br>(0.60-1.08) | 0.86<br>(0.63-1.15) | 0.53<br>(0.28-1.01) | 0.64<br>(0.33-1.22) | **1.11**<br>**(1.02-1.20)** | **1.14**<br>**(1.05-1.24)** |
| Craniosacral Therapy | 0.82<br>(0.31-2.14) | 0.84<br>(0.31-2.30) | NE | NE | NE | NE | NE | NE | NE | NE | NE | NE | 0.85<br>(0.32-2.22) | 0.85<br>(0.31-2.36) |
| Energy Healing Therapy | 1.39<br>(0.98-1.96) | 1.32<br>(0.95-1.83) | **1.90**<br>**(1.41-2.57)** | **2.32**<br>**(1.48-3.65)** | NE | NE | NE | NE | 0.89<br>(0.21-3.80) | 0.90<br>(0.24-3.41) | NE | NE | 1.24<br>(0.76-2.00) | 1.13<br>(0.73-1.74) |
| Herbal Supplements <sup>c, d</sup> | **0.33**<br>**(0.29-0.37)** | **0.34**<br>**(0.30-0.38)** | **0.46**<br>**(0.36-0.60)** | **0.47**<br>**(0.36-0.61)** | 0.15<br>(0.01-1.67) | 0.08<br>(0.00-2.32) | **0.36**<br>**(0.22-0.59)** | **0.37**<br>**(0.23-0.59)** | **0.50**<br>**(0.39-0.65)** | **0.50**<br>**(0.38-0.65)** | 0.70<br>(0.43-1.15) | 0.66<br>(0.40-1.10) | **0.28**<br>**(0.24-0.33)** | **0.29**<br>**(0.25-0.34)** |
| Homeopathy | 0.98<br>(0.77-1.24) | 0.89<br>(0.70-1.13) | 0.93<br>(0.50-1.73) | 0.88<br>(0.44-1.75) | NE | NE | 0.91<br>(0.30-2.78) | 0.95<br>(0.31-2.92) | 0.67<br>(0.27-1.67) | 0.60<br>(0.23-1.54) | 1.06<br>(0.41-2.76) | 0.78<br>(0.32-1.89) | 1.01<br>(0.77-1.33) | 0.92<br>(0.68-1.23) |
| Hypnosis | 0.89<br>(0.54-1.47) | 0.91<br>(0.54-1.54) | NE | NE | NE | NE | NE | NE | NE | NE | NE | NE | 0.99<br>(0.60-1.64) | 1.03<br>(0.62-1.71) |
| Massage | **1.40**<br>**(1.29-1.51)** | **1.39**<br>**(1.29-1.50)** | **1.27**<br>**(1.04-1.56)** | **1.25**<br>**(1.03-1.52)** | 0.97<br>(0.23-4.11) | 1.56<br>(0.32-7.55) | **1.56**<br>**(1.22-1.99)** | **1.45**<br>**(1.11-1.89)** | **1.44**<br>**(1.16-1.80)** | **1.42**<br>**(1.13-1.79)** | 1.47<br>(0.92-2.35) | **1.69**<br>**(1.02-2.80)** | **1.41**<br>**(1.28-1.54)** | **1.39**<br>**(1.26-1.53)** |
| Meditation, Guided Imagery, or<br>Progressive relaxation | **1.90**<br>**(1.76-2.04)** | **1.82**<br>**(1.69-1.97)** | **1.73**<br>**(1.45-2.06)** | **1.66**<br>**(1.37-2.00)** | 1.56<br>(0.76-3.20) | 1.73<br>(0.64-4.65) | **1.80**<br>**(1.33-2.43)** | **1.81**<br>**(1.33-2.47)** | **2.05**<br>**(1.76-2.38)** | **1.99**<br>**(1.70-2.32)** | 1.46<br>(0.94-2.28) | 1.23<br>(0.81-1.88) | **1.92**<br>**(1.75-2.11)** | **1.83**<br>**(1.65-2.02)** |
| Movement or exercise techniques | **1.71**<br>**(1.47-1.99)** | **1.55**<br>**(1.33-1.80)** | **1.94**<br>**(1.58-2.38)** | **1.73**<br>**(1.34-2.23)** | NE | NE | 1.19<br>(0.51-2.76) | 1.33<br>(0.59-3.01) | 1.50<br>(0.99-2.26) | **1.64**<br>**(1.03-2.63)** | NE | NE | **1.73**<br>**(1.43-2.08)** | **1.58**<br>**(1.30-1.91)** |
| Naturopathy | **1.39**<br>**(1.01-1.90)** | 1.29<br>(0.93-1.80) | 1.68<br>(0.96-2.94) | 2.14<br>(0.92-4.98) | NE | NE | NE | NE | **1.72**<br>**(1.05-2.82)** | **1.56**<br>**(1.06-2.31)** | 1.34<br>(0.42-4.28) | 1.24<br>(0.39-3.95) | 1.30<br>(0.87-1.95) | 1.16<br>(0.76-1.76) |
| Special diets | 1.08<br>(0.91-1.29) | 1.08<br>(0.90-1.29) | 1.17<br>(0.81-1.68) | 1.20<br>(0.80-1.79) | NE | NE | 0.96<br>(0.57-1.60) | 1.05<br>(0.61-1.80) | 0.90<br>(0.58-1.39) | 0.88<br>(0.57-1.34) | 0.66<br>(0.19-2.26) | 0.70<br>(0.25-1.93) | 1.11<br>(0.88-1.41) | 1.13<br>(0.89-1.42) |
| Traditional Healers <sup>d</sup> | 0.94<br>(0.67-1.32) | 0.81<br>(0.56-1.17) | 0.82<br>(0.52-1.28) | 0.91<br>(0.58-1.43) | 0.65<br>(0.24-1.79) | 0.45<br>(0.19-1.10) | NE | NE | 0.50<br>(0.06-4.33) | 0.39<br>(0.04-3.86) | 0.34<br>(0.03-3.58) | 0.46<br>(0.03-6.43) | 1.37<br>(0.82-2.29) | 1.38<br>(0.77-2.48) |
| Yoga, Tai Chi, or Qigong | **1.51**<br>**(1.41-1.62)** | **1.40**<br>**(1.30-1.51)** | **1.51**<br>**(1.26-1.80)** | **1.38**<br>**(1.15-1.66)** | NE | NE | **1.79**<br>**(1.40-2.27)** | **1.80**<br>**(1.41-2.30)** | 1.20<br>(0.93-1.56) | 1.18<br>(0.91-1.53) | **1.65**<br>**(1.13-2.41)** | **1.84**<br>**(1.27-2.65)** | **1.51**<br>**(1.38-1.64)** | **1.37**<br>**(1.25-1.50)** |

Abbreviations: CAM (Complementary and Alternative Medicine), NH (non-Hispanic), NE (Not able to estimate)
<sup>a</sup> Models adjusted for age (years), sex (men, women), race and ethnicity (Hispanic/Latine, NH-American Indian/Alaska Native, NH-Asian, NH-Black/African American, NH-Multiple race or other group, NH-White) when not stratified by race and ethnicity, educational attainment (≤high school, some college, ≥college), family income (<\$35,000, \$35,000-\$74,999, ≥\$75,000), employment status (employed (≥ 35 hours), employed (< 35 hours), not employed/not in labor force), marital status (divorced/widowed/separated/married, spouse absent, Single/Never married, married, spouse present/living with partner), body mass index (underweight/recommended (<25 kg/m<sup>2</sup>), overweight (25-30 kg/m<sup>2</sup>), obesity (≥30 kg/m<sup>2</sup>)), general health status (excellent, very good, good, fair/poor), and region of residence (Northwest, Midwest, South, West)
<sup>b</sup> CAM therapy most important to respondent's health (Yes vs No)
<sup>c</sup> Includes combinations of herbal supplements
<sup>d</sup> Significant interaction effect between race and ethnicity and CAM therapy most important to respondent's health on perceived better sleep (p < .05)
\* All estimates are weighted for the complex survey design. Bolded values indicate statistical significance at a two-sided p-value < 0.05

#### By race and ethnicity

Based on Wald tests (all p <0.05), race and ethnicity modified associations between use/prioritization of chiropractic or osteopathic manipulation, herbal supplements, and traditional healers with perceiving sleep benefits of those therapies. Among Asian adults, participants who reported versus did not report chiropractic or osteopathic manipulation as most important for their health were 48% less likely (aPR=0.52 [95% CI:0.31-0.87]) to report that therapy helped them achieve ‘better sleep’. Conversely, prioritizing chiropractic or osteopathic manipulation was associated with perceiving ‘better sleep’ among NH-White adults (aPR=1.14 [95% CI:1.05-1.24]) and was not associated with perceiving ‘better sleep’ among Hispanic/Latine, NH-AI/AN, or NH-Black adults. However, the small sample size among NH-AI/AN adults resulted in imprecise estimates that should be interpreted with caution. Across groups, adults who reported versus did not report herbal supplements as most important for their health were less likely to report that herbal supplements helped them achieve better sleep; however, magnitudes of associations varied by race and ethnicity with the largest estimates among NH-White adults. Herbal supplements deemed most important for health were associated with a 71% lower prevalence (aPR=0.29 [95% CI:0.25-0.34]) of perceiving ‘better sleep’ among NH-White adults; CIs overlapped with those among NH-Asian adults (aPR=0.37 [95% CI:0.23-0.59]); and associations were weaker among Hispanic/Latine (aPR=0.47 [95% CI:0.36-0.61]) and NH-Black (aPR=0.50 [95% CI:0.38-0.65]) adults. Despite Wald tests suggesting effect modification by race and ethnicity for associations between traditional healers and perceiving ‘better sleep’, estimates were imprecise across groups – likely related to low prevalence of reports of use/health benefits in most groups, reducing interpretability.

## DISCUSSION

In this nationally representative sample of U.S. adults who reported using CAM, associations between CAM approaches considered most important for health and perceived sleep benefits varied by race and ethnicity for certain modalities. Perceived sleep benefits from mind-body (i.e., meditation, guided imagery, progressive relaxation), soft tissue and movement (i.e., massage; movement or exercise techniques), and hybrid meditative/movement practices (i.e., yoga, Tai Chi, Qigong) were similar across racial and ethnic groups, while sleep benefits of other CAM approaches varied. Specifically, musculoskeletal manipulation (i.e., chiropractic or osteopathic manipulation) was associated with perceiving sleep benefits only among NH-White adults; conversely, musculoskeletal manipulation was associated with lower likelihood of perceived sleep benefits among NH-Asian adults. Herbal supplements were the most reported CAM modality, overall; however, they were associated with lower prevalence of perceiving ‘better sleep’, with the largest magnitude of association among NH-White adults. Our findings did not support our hypothesis that individual CAM therapies are more strongly associated with better sleep among racial and ethnic groups other than NH-White adults. Other CAM modalities were generally not associated with perceived sleep benefits, suggesting that specific healing practices, cultural context, and health beliefs may influence how individuals experience and evaluate CAM therapies in relation to sleep.

Despite limited comparisons to prior studies, the directions of the observed racial and ethnic differences in relationships between chiropractic or osteopathic manipulation and herbal supplement use with perceived sleep benefits are plausible. Specifically, our results among NH-White adults align with a prior descriptive study using 2012 NHIS data that similarly reported an association between spinal manipulation and sleep benefits (34); however, racial and ethnic differences were not examined. No identified international or U.S. studies assessed these associations by race and ethnicity. Although a prior study reported fewer perceived sleep health benefits with any CAM use among NH-White adults, it was not specific to individual CAM approaches and focused only on U.S. adults aged 50 years and older (31). Our observation of adults commonly considering herbal supplements as most important for health, yet its association with a lower prevalence of better sleep is consistent with prior literature. The 2012 NHIS study reported that ∼89% adults reported using natural product supplements for wellness; however, only 21% reported perceived sleep improvements with use (34). This was notably lower than perceived sleep improvements with other CAM modalities such as spinal manipulation and hybrid meditative/movement practices (34). However, no international or U.S. studies investigated this association by race and ethnicity, limiting comparisons to our findings of the strongest associations with not perceiving sleep benefits among NH-White adults. Extending the literature, we identified that perceived benefits or lack thereof may not be uniform across racial and ethnic groups, although replication is needed.

Racial and ethnic differences in perceived sleep benefits from musculoskeletal manipulation and herbal remedies may reflect variation in use, familiarity, and cultural acceptance. NH-White adults are more likely to use both modalities than other racial and ethnic groups –particularly NH-Asian adults for chiropractic care (38)– which may contribute to differences in perceived benefits across groups (29, 38-40). Further, barriers to CAM access (14), racial differences in reporting sleep complaints (7), and heterogenous racial and ethnic categories (e.g., NH-Asian) (41) may contribute to observations. Overall, herbal supplements are frequently reported as being used for general wellness; however, sleep benefits may not be expected (34, 42). Further, broad use of these supplements for non-sleep related reasons are more prevalent among NH-White adults and may contribute to stronger negative associations with ‘better sleep’ observed among NH-White adults (43). Additionally, evidence regarding the effectiveness of herbal supplements for insomnia and sleep disturbances are mixed (44-46), potentially reflecting variation in herbal supplement composition, dosage, quality, and regulation as well as sleep dimension (47-49). Moreover, caffeine-containing supplements (e.g., green tea, energy supplements) can impair sleep if taken later in the day (50). Combining diverse herbal supplements in this study may dilute or mask perceived sleep benefits. Future research should assess more specific herbal supplements and identify factors underlying racial and ethnic differences in perceived sleep benefits from specific CAM therapies.

In contrast, mind-body and body-based therapies may address behavioral, emotional, and physical factors that influence sleep across populations. Meditation, guided imagery, progressive relaxation, and massage were consistently associated with better sleep across racial and ethnic groups, aligning with evidence that mind-body therapies may improve sleep quality and insomnia symptoms (18, 21, 26). Meditation and relaxation practices are hypothesized to improve sleep by reducing cognitive and physiological arousal, stress reactivity, and rumination, all of which interfere with sleep initiation and maintenance (20, 51). Similarly, massage may improve sleep by reducing pain, muscle tension, physical discomfort, and sympathetic nervous system activation (26, 52). While mindfulness-based interventions are well-studied in sleep literature, fewer population-based studies have examined massage as a sleep-promoting therapy. The consistent associations across the overall population suggest that massage may be an underrecognized approach for supporting sleep health.

Movement-based and hybrid meditative/movement therapies also demonstrated associations with perceived sleep improvement across all adults, consistent with previous research demonstrating sleep and overall well-being benefits (19, 44, 53). Movement-based approaches may improve sleep through increased physical activity, improved mood, reduced stress, and enhanced self- and autonomic regulation (54). Notably, yoga, Tai Chi, and Qigong practices originated in East and South Asian healing traditions emphasizing balance, mindful movement, breath regulation, and holistic wellbeing (53, 55). Therefore, these associations may reflect stronger alignment with whole-person or interconnected dimensions of health (56), which has potentially positive implications for sleep (16, 25).

Among NH-AI/AN participants, 31% identified traditional healers as their most important CAM modality, a prevalence higher than other racial or ethnic groups, highlighting the importance of Indigenous healing traditions and culturally grounded approaches to wellness. Although traditional healers were not associated with perceiving ‘better sleep’, null results are likely explained by imprecise estimates due to the small sample size and limited statistical power, and potentially, by the broader goals of traditional healing, which emphasize spiritual, cultural, and community wellbeing rather than sleep (57). Future studies incorporating culturally relevant measures of wellbeing may provide a better understanding of how traditional healing approaches influence sleep and health overall. Additionally, studies are needed on other CAM modalities that were not associated with perceived sleep benefits in this study, such as acupuncture, which has previously been associated with reduced insomnia severity (58). Given the limited, low-certainty evidence, these therapies warrant further investigation.

Several limitations and strengths should be considered. The cross-sectional design precludes establishing temporality and causality. Also, all measures were self-reported and susceptible to misclassification, including ‘better sleep’. Nonetheless, the construct may reflect subjective experiences of relaxation, wellbeing, or restoration not captured by other survey measures. Further, perception or subjective appraisal is important for health outcomes (59, 60). Participants identified only the CAM therapy most important to their health, which may underestimate the use of multiple therapies simultaneously, and the entire sample reported use of a CAM therapy. Use of other therapies may impact results. Moreover, results are not generalizable to populations who report no CAM therapy use. Additionally, sample sizes for some groups (e.g., NH-AI/AN and multiracial) were small, limiting statistical precision and interpretability for some estimates. However, strengths include use of a large nationally representative sample, examination of numerous CAM modalities, and ability to examine these associations across multiple racial and ethnic groups.

Overall, perceived importance of CAM for health and its association with perceiving sleep benefits varied across racial and ethnic groups for certain modalities. While mind-body, soft tissue and movement techniques, along with hybrid meditative/movement practices were associated with perceived sleep benefits across all groups, musculoskeletal manipulation was beneficial only among NH-White adults – the group in which herbal supplement use was most strongly reported as not beneficial for sleep. These findings highlight the importance of considering specific CAM modalities within broader, person-centered (or directed) discussions of sleep health to ensure modalities match the specific need and suggest cultural context may influence perceived benefits. Given CAM’s holistic approach, future interventions may benefit from integrating evidence-based behavioral sleep strategies with culturally meaningful wellness practices to enhance engagement, acceptability, and perceived sleep benefits among diverse populations.

## Data Availability

The datasets analyzed during the current study are publicly available from the National Center for Health Statistics (NCHS), Centers for Disease Control and Prevention, National Health Interview Survey (NHIS), at: https://www.cdc.gov/nchs/nhis/index.htm

## FUNDING

This research was supported in part by the Intramural Research Program of the National Institutes of Health (NIH), National Institute of Environmental Health Sciences (Z1AES103325 [CLJ]). The contributions of the NIH authors are considered Works of the United States Government. The findings and conclusions presented in this paper are those of the authors and do not necessarily reflect the views of the NIH or the U.S. Department of Health and Human Services.

## ACKNOWLEDGEMENTS

The authors would like to thank all respondents who participated in the NHIS survey. The authors also would like to thank Kennedy D.R. Williams for assistance with reviewing the prior literature.

## AUTHOR CONTRIBUTIONS

*Authors:* Rupsha Singh, Symielle A. Gaston, Christopher Payne, Dayna T. Neo, Suzanne M. Bertisch, Chandra L. Jackson

*Study concept and design:* CL. Jackson.

*Acquisition of data:* Christopher Payne, CL. Jackson.

*Statistical Analysis:* Christopher Payne.

*Interpretation of data:* Rupsha Singh, Symielle A. Gaston, Christopher Payne, Dayna T. Neo, Suzanne M. Bertisch, Chandra L. Jackson.

*Drafting of the manuscript:* Rupsha Singh, Symielle A. Gaston, Christopher Payne, Dayna T. Neo.

*Critical revision of the manuscript for important intellectual content:* Rupsha Singh, Symielle A. Gaston, Christopher Payne, Dayna T. Neo, Suzanne M. Bertisch, Chandra L. Jackson.

*Administrative, technical, and material support:* CL. Jackson.

*Obtaining funding and study supervision:* CL. Jackson.

*Final Approval:* Rupsha Singh, Symielle A. Gaston, Christopher Payne, Dayna T. Neo, Suzanne M. Bertisch, Chandra L. Jackson.

## DECLARATION OF CONFLICTS OF INTEREST

The authors declare no conflict of interest.

## DECLARATION OF GENERATIVE AI and AI-ASSISTED TECHNOLOLGIES

During the preparation of this work, the authors wrote the manuscript text independently and subsequently used Copilot to edit and refine the manuscript text. After using this tool, the authors reviewed and edited the content as needed and take full responsibility for the content of the publication.

## Notes

### Competing Interest Statement

The authors have declared no competing interest.

