## Supplemental Material for "Complementary and Alternative Medicine Therapies in relation to Self-Reported Better Sleep across Racial and Ethnic Groups in the United States"

Rupsha Singh, PhD, DBSM ^1^*

Symielle A. Gaston, PhD, MPH ^1^*

Christopher Payne, MA ^2^

Dayna T. Neo, PhD, MPH ^2^

Suzanne M. Bertisch, MD, MPH ^3^

Wayne B. Jonas, MD ^4^

Chandra L. Jackson, PhD, MS ^1, 5^

^1^ Epidemiology Branch, National Institute of Environmental Health Sciences, National Institutes of Health, Department of Health and Human Services, Research Triangle Park, NC, USA

^2^ DLH, LLC, Bethesda, MD, USA

^3^ Division of Sleep and Circadian Disorders, Brigham and Women's Hospital, Harvard Medical School, Boston, MA, USA

^4^ Healing Works Foundation and Georgetown University School of Medicine, Washington, D.C., USA

^5^ Intramural Program, National Institute on Minority Health and Health Disparities, National Institutes of Health, Department of Health and Human Services, Bethesda, MD, USA

* Equal contributions

Please direct correspondence to Dr. Chandra L. Jackson at 111 TW Alexander Drive, MD A3-05, Research Triangle Park, N.C. 27709; telephone: 984-287-3701; fax: 301-480-3290;.

### Supplemental Figure 1. Flow chart of study population selection

Final analytic sample

**(N=9,308)**

Total NHIS Sample,

2012

**(N=47,800)**

- Missing data
  - Better sleep (n=177)
  - Sleep duration (n=29)
  - Insomnia (n=1)
  - Age (n=0)
  - Sex (n=0)
  - Race and Ethnicity (n=18)
  - Educational Attainment (n=18)
  - Annual household income (n=431)
  - Employment/Work status (n=2)
  - Marital status (n=10)
  - Body mass index (n=185)
  - General health status (n=2)
  - Region of residence (n=0)
- **Total excluded (n=873)**
- Sample adults who did not give responses to any of the questions in the supplement (n=931)
- No CAM therapy use (n=23,413)

NHIS Sample of adults reported using CAM at least once in the past year, 2012

**(N=10,181)**

Participants with sample adult

interview data

**(N=34,525)**

- Sample child (age <18 years) data (n=13,275)

### Supplemental Table 1. Comparison of included and excluded study participants, National Health Interview Survey, 2012, (N=10,181)

|  | **Included**  n=9,308 (91.4%) | **Excluded**  n=873 (8.6%) | Chi-square or  t-test p-value |
| --- | --- | --- | --- |
| Sociodemographic Characteristics* |  |  |  |
| Age (years), mean (SE) | 46.4 (0.3) | 53.0 (0.8) | <0.001 |
| Age group |  |  | <0.001 |
| 18-30 years | 21.2 | 10.8 |  |
| 31-49 years | 35.1 | 27.3 |  |
| ≥ 50 years | 43.6 | 62.0 |  |
| Sex |  |  | <0.001 |
| Men | 41.8 | 31.6 |  |
| Women | 58.2 | 68.4 |  |
| Race/ethnicity |  |  | 0.028 |
| Hispanic/Latine | 9.7 | 7.3 |  |
| NH-American Indian/Native Alaskan | 0.5 | 0.3 |  |
| NH-Asian | 5.4 | 5.5 |  |
| NH-Black/African American | 6.2 | 8.1 |  |
| NH-multiple race groups | 2.0 | 0.9 |  |
| NH-White | 76.4 | 78.0 |  |
| Educational Attainment |  |  | 0.038 |
| <High School | 6.3 | 6.5 |  |
| High School graduate | 18.7 | 23.7 |  |
| Some College | 33.9 | 32.4 |  |
| ≥College | 41.1 | 37.4 |  |
| Annual household income |  |  | 0.265 |
| < $35,000 | 24.4 | 27.4 |  |
| $35,000-$74,999 | 31.5 | 33.5 |  |
| ≥ $75,000 | 44.1 | 39.1 |  |
| Employment/Work status |  |  | <0.001 |
| Employed (≥ 35 hours) | 54.3 | 43.2 |  |
| Employed (< 35 hours) | 13.0 | 12.2 |  |
| Not employed/not in labor force | 32.7 | 44.6 |  |
| Marital status |  |  | 0.021 |
| Divorced/Widowed/Separated/Married, spouse absent | 17.1 | 19.3 |  |
| Single/Never married | 19.7 | 15.1 |  |
| Married, spouse present/living with partner | 63.2 | 65.7 |  |
| Region of residence |  |  | <0.001 |
| Northeast | 16.3 | 24.0 |  |
| Midwest | 25.5 | 26.7 |  |
| South | 29.3 | 23.2 |  |
| West | 28.9 | 26.1 |  |
| Health Behaviors* |  |  |  |
| Usual sleep duration |  |  | 0.380 |
| Short (<7 hours) | 30.6 | 29.7 |  |
| Recommended (7-9 hours) | 66.5 | 66.4 |  |
| Long (>9 hours) | 2.9 | 4.0 |  |
| Insomnia (Yes) | 24.4 | 25.8 | 0.467 |
| Smoking status |  |  | 0.495 |
| Never/quit >12 months prior | 83.5 | 85.5 |  |
| Former/quit≤12 months ago | 2.1 | 2.1 |  |
| Current | 14.4 | 12.4 |  |
| Alcohol consumption |  |  | 0.008 |
| Current (≥1 drink past year) | 74.4 | 70.0 |  |
| Former (no drinks past year) | 13.2 | 12.8 |  |
| Lifetime abstinence (<12 drinks in life) | 12.4 | 17.2 |  |
| Leisure-time physical activity (PA) |  |  | 0.030 |
| Never/unable | 19.7 | 24.4 |  |
| Does not meet PA guidelines | 18.2 | 18.7 |  |
| Meets PA guidelines ^a^ | 62.1 | 56.8 |  |
| Clinical Characteristics* |  |  |  |
| General health status |  |  | 0.205 |
| Excellent | 30.6 | 28.5 |  |
| Very good | 34.9 | 34.1 |  |
| Good | 24.5 | 24.5 |  |
| Fair/poor | 10.0 | 12.9 |  |
| Body mass index category |  |  | 0.225 |
| Underweight (<18.5 kg/m^2^) | 1.8 | 1.2 |  |
| Recommended (18.5-<25 kg/m^2^) | 38.5 | 38.8 |  |
| Overweight (25-<30 kg/m^2^) | 34.1 | 37.7 |  |
| Obesity (≥30 kg/m^2^) | 25.6 | 22.3 |  |
| Dyslipidemia (Yes) ^b^ | 19.3 | 23.7 | 0.011 |
| Hypertension (Yes) ^c^ | 19.7 | 26.0 | 0.001 |
| Prediabetes/Diabetes (Yes) ^d^ | 13.0 | 18.5 | 0.001 |
| SPD (Yes) ^e^ | 3.1 | 4.0 | 0.385 |
| Abbreviations: CAM = Complementary and Alternative Medicine, NH = non-Hispanic, PA = physical activity, SPD = Serious psychological distress, SE = standard error | | | |
| Note: Data are presented as column percentages or means and standard errors. Percentages may not sum to 100 due to missing or rounding.  * All estimates are weighted for the survey's complex sampling design. | | | |
| ^a^ Meets PA guidelines is defined as ≥150 minutes/week of moderate intensity or ≥75 minutes/week of vigorous intensity or ≥150 minutes/week of moderate and vigorous intensity | | | |
| ^b^ Dyslipidemia defined as had high cholesterol during the past 12 months. | | | |
| ^c^ Hypertension defined as had hypertension or high blood pressure during the past 12 months. | | | |
| ^d^ Prediabetes defined as ever told by a doctor or other health professional they have prediabetes, impaired fasting glucose, impaired glucose tolerance, borderline diabetes, or high blood sugar. Diabetes defined as ever told by a doctor or other health professional that they have diabetes or sugar diabetes. | | | |
| ^e^ Serious psychological distress defined as a score ≥13 on the Kessler-6 Psychological Distress Scale | | | |

### Supplemental Table 2. Weighted prevalence of self-reported better sleep with the use by modality of Complementary and Alternative Medicine therapy, overall and by race and ethnicity, National Health Interview Survey, 2012 (N=9,308)

|  | **Overall**  **n=9,308 (100%)** | **Hispanic/Latine n=1,071 (11.5%)** | **NH-American Indian/**  **Alaska Native**  **n=55 (0.6%)** | **NH-Asian**  **n=616 (6.6%)** | **NH-Black/**  **African American**  **n=770 (8.3%)** | **NH-Multiple race groups**  **n=213 (2.3%)** | **NH-White**  **n=6,583 (70.7%)** |
| --- | --- | --- | --- | --- | --- | --- | --- |
| CAM Therapies* | | | | | | | |
| Overall | 40.6 | 46.3 | 55.0 | 41.3 | 46.0 | 45.5 | 39.2 |
| Acupuncture | 38.6 | 41.2 | 0.0 | 34.2 | 39.4 | 0.0 | 39.6 |
| Biofeedback | 39.5 | 51.9 | 0.0 | 0.0 | 71.8 | 0.0 | 37.4 |
| Chiropractic and Osteopathic Manipulation | 41.5 | 42.6 | 67.4 | 23.8 | 38.3 | 26.4 | 42.1 |
| Craniosacral Therapy | 33.2 | 0.0 | 0.0 | 0.0 | 0.0 | 0.0 | 33.2 |
| Energy Healing Therapy | 56.2 | 87.7 | 0.0 | 0.0 | 41.1 | 100.0 | 48.4 |
| Herbal Supplements ^a^ | 16.5 | 25.1 | 9.2 | 18.4 | 27.8 | 34.9 | 13.8 |
| Homeopathy | 39.8 | 43.4 | 0.0 | 37.8 | 30.9 | 48.0 | 39.7 |
| Hypnosis | 36.2 | 0.0 | 0.0 | 0.0 | 0.0 | 0.0 | 39.0 |
| Massage | 54.0 | 56.7 | 53.2 | 59.7 | 62.8 | 64.8 | 52.4 |
| Meditation, guided imagery, or progressive relaxation | 72.6 | 76.5 | 80.5 | 70.8 | 86.3 | 63.6 | 71.1 |
| Movement or exercise techniques | 68.7 | 88.7 | 0.0 | 49.0 | 68.1 | 100.0 | 67.0 |
| Naturopathy | 56.4 | 77.9 | 0.0 | 100.0 | 78.8 | 60.7 | 50.9 |
| Special diets | 43.8 | 53.7 | 100.0 | 39.6 | 41.4 | 30.9 | 43.5 |
| Traditional Healers | 38.2 | 38.3 | 40.2 | 0.0 | 23.2 | 15.8 | 53.8 |
| Yoga, Tai Chi, or Qigong | 57.5 | 65.2 | 100.0 | 62.7 | 53.6 | 66.7 | 55.6 |
| Abbreviations: CAM = Complementary and Alternative Medicine, NH = non-Hispanic  Note: Data presented are the sample proportions of participants who reported ‘better sleep’ as a result of using specific CAM therapies that were also identified by the participant as most important for their health. | | | | | | | |
| * All estimates are weighted for the survey's complex sampling design. | | | | | | | |
| ^a^ Includes combinations of herbal supplements | | | | | | | |

### Appendix I – CAM therapies

*Acupuncture*

The 2012 field representative’s manual defines acupuncture as a family of procedures involving stimulation of anatomical points on the body by a variety of techniques. American practices of acupuncture incorporate medical traditions from China, Japan, Korea, and other countries. The acupuncture technique most studied scientifically involves penetrating the skin with thin, solid, metallic needles that are manipulated by hand or by electrical stimulation.

*Ayurveda*

The 2012 field representative’s manual defines ayurveda as a system of medicine that originated in India several thousand years ago. In the United States, Ayurveda is considered a type of CAM and a whole medical system. As with other such systems, it is based on theories of health and illness and on ways to prevent, manage, or treat health problems. Ayurveda aims to integrate and balance the body, mind, and spirit (thus, some view it as "holistic"). This balance is believed to lead to contentment and health and to help prevent illness. However, Ayurveda also proposes treatments for specific health problems, whether they are physical or mental. An aim of Ayurvedic practices is to cleanse the body of substances that can cause disease, and this is believed to help reestablish harmony and balance.

*Biofeedback*

The 2012 field representative’s manual defines biofeedback as a therapy that uses simple electronic devices to teach clients how to consciously regulate bodily functions, such as breathing, heart rate, and blood pressure, to improve overall health. Biofeedback is used to reduce stress, eliminate headaches, recondition injured muscles, control asthmatic attacks, and relieve pain.

*Chelation therapy*

The 2012 field representative’s manual defines chelation therapy as a chemical process in which a substance is used to bind molecules, such as metals or minerals, and hold them tightly so that they can be removed from a system, such as the body. In medicine, chelation has been scientifically proven to rid the body of excess or toxic metals. For example, a person who has lead poisoning may be given chelation therapy to bind and remove excess lead from the body before it can cause damage.

*Chiropractic or osteopathic manipulation*

The 2012 field representative’s manual defines chiropractic manipulation as a form of health care that focuses on the relationship between the body's structure, primarily of the spine, and function. Chiropractors or chiropractic physicians use a type of hands-on therapy called manipulation (or adjustment) as their core clinical procedure.

The 2012 field representative’s manual defines osteopathic manipulation as a full-body system of hands-on techniques to alleviate pain, restore function, and promote health and well-being.

*Craniosacral therapy*

The 2012 field representative’s manual defines craniosacral therapy as a body-based practice. Practitioners use light touch and manipulation focused on the skull and spine, with the intent of sensing and removing what they refer to as blockages or imbalances that may be contributing to a health condition.

*Energy healing therapy*

The 2012 field representative’s manual defines energy healing therapy as the channeling of healing energy through the hands of a practitioner into the client's body to restore a normal energy balance and, therefore, health. Energy healing therapy has been used to treat a wide variety of ailments and health problems and is often used in conjunction with other alternative and conventional medical treatments.

*Homeopathy*

The 2012 field representative’s manual defines homeopathy as a system of medical practices based on the theory that any substance that can produce symptoms of disease or illness in a healthy person can cure those symptoms in a sick person. For example, someone suffering from insomnia may be given a homeopathic dose of coffee. Administered in diluted form, homeopathic remedies are derived from many natural sources, including plants, animals, metals, and minerals.

*Hypnosis*

The 2012 field representative’s manual defines hypnosis as an altered state of consciousness characterized by increased responsiveness to suggestion. This hypnotic state is attained by first relaxing the body, then shifting attention toward a narrow range of objects or ideas as suggested by the hypnotist or hypnotherapist. The procedure is used to effect positive changes and to treat numerous health conditions including ulcers, chronic pain, respiratory ailments, stress, and headaches.

*Massage therapy*

The 2012 field representative’s manual defines massage therapy as the manipulation of muscle and connective tissue to enhance function of those tissues and promote relaxation and well-being.

*Naturopathy*

The 2012 field representative’s manual defines naturopathy as an alternative medical system. Naturopathic medicine proposes that there is a healing power in the body that establishes, maintains, and restores health. Practitioners work with the patient with a goal of supporting this power through treatments such as nutrition and lifestyle counseling, dietary supplements, medicinal plants, exercise, homeopathy, and treatments from traditional Chinese medicine.

*Yoga/Tai-chi/Qigong*

The 2012 field representative’s manual defines yoga as a technique that combines breathing exercises, physical postures, and meditation to calm the nervous system and balance body, mind, and spirit. Usually performed in classes, sessions are conducted once a week or more and roughly last 45 minutes.

The 2012 field representative’s manual defines Tai-chi as a mind-body practice that originated in China as a martial art. A person doing tai-chi moves his body slowly and gently, while breathing deeply and meditating (tai-chi is sometimes called "moving meditation"). Many practitioners believe that tai-chi helps the flow throughout the body of a proposed vital energy called “qi.” A person practicing tai-chi moves her body in a slow, relaxed, and graceful series of movements. One can practice on one's own or in a group. The movements make up what are called forms (or routines).

The 2012 field representative’s manual defines Qigong as an ancient Chinese discipline combining the use of gentle physical movements, mental focus, and deep breathing directed toward specific parts of the body. Performed in repetitions, the exercises are normally performed two or more times a week for 30 minutes at a time.

### Appendix II – Traditional healers

*Curandero, Machi or Parchero*

The 2007 and later field representative's manuals state that a Curandero, Machi or Parchero are a type of traditional folk healer. Originally found in Latin America, Curanderos specialize in treating illness through the use of supernatural forces, herbal remedies, and other natural medicines. "Machi" most commonly refers to a traditional healer, shaman, and spiritual leader within the Mapuche culture of Chile and Argentina. These individuals are respected figures with deep knowledge of medicinal herbs, dream interpretation, and spiritual rituals.

*Hierbero, Yerbero or Hierbista*

The 2007 and later field representative's manuals define a Hierbo, Yerbera, or (starting in 2012) a Hierbista or Yerbero as a practitioner with knowledge of the medicinal qualities of plants.

*Huesero*

The 2012 field representative’s manual states that a Huesero or “bone setter” (in Hispanic folk healing), specializes in bone ailments, mainly lesions and fractures.

*Native American healers/Medicine men*

The 2007 and later field representative's manuals state that Native American healers/Medicine men use information from the "spirit world" to benefit the community. People see Native American healers for a variety of reasons, especially to find relief or a cure from illness or to find spiritual guidance.

*Shaman*

The 2007 and later field representative's manuals state that Shamans are said to act as mediums between the invisible spiritual world and the physical world. Most gain knowledge through contact with the spiritual world and use the information to perform tasks such as divination, influencing natural events, and healing the sick or injured.

*Sobador*

The 2007 and later field representative's manuals state that a Sobador uses massage and rub techniques to treat patients.

### Appendix III – Herbal and non-vitamin supplements

Acai (pills, gelcaps),
Bee Pollen and other Bee products,
Chondroitin,
Co-enzyme Q10 (CoQ10),
Cranberry (pills or capsules),
Digestive Enzymes (lactaid),
Echinacea,
Fish Oil or omega 3 or DHA fatty acid or EPA fatty acid supplements,
Garlic supplements (pills, gelcaps),
Ginkgo Biloba,
Ginseng,
Glucosamine,
Green tea pills (not brewed tea) or EGCG (pills),
Melatonin,
Milk Thistle (silymarin),
MSM (Methylsulfonylmethane),
Probiotics or Prebiotics,
SAM-e,
Saw Palmetto,
Valerian

### Appendix IV – Meditation, guided imagery, and progressive relaxation techniques

The 2012 field representative’s manual defines meditation as a group of techniques, most of which started in eastern religious or spiritual traditions. In meditation, a person learns to focus their attention and suspend the stream of thoughts that normally occupy the mind. This practice is believed to result in a state of greater physical relaxation, mental calmness, and psychological balance. Practicing meditation can change how a person relates to the flow of emotions and thoughts in the mind. In mantra meditation, the meditator focuses on a mantra (a specially chosen word, sound, or phrase repeated silently). Mindfulness meditation is a type of meditation based on the concept of being mindful, or having increased awareness, of the present. It uses breathing methods, guided imagery, and other practices to relax the body and mind and help reduce stress. It is also known as mindfulness relaxation and mindfulness-based stress reduction. Spiritual meditation may be performed according to the practices of one of the major religions or within a spiritual tradition. The techniques used may be the same as in other types of meditation (for example, transcendental meditation), but the focus is on spirituality (such as repeating a spiritual, meditative phrase).

The 2012 field representative’s manual states that guided imagery is used for healing or health maintenance and involves a series of relaxation techniques followed by the visualization of detailed images, usually calm and peaceful in nature. If used for treatment, the individual will visualize their body free of the specific problem or condition. Sessions are typically 20 to 30 minutes long and may be practiced several times a week.

The 2012 field representative’s manual states that progressive relaxation is used to relieve tension and stress by systematically tensing and relaxing successive muscle groups.

### Appendix V – Movement and exercise techniques

*Alexander technique*

The 2012 field representative’s manual defines the Alexander technique as a practice that uses guidance and education on ways to improve posture and movement. The intent is to teach a person how to use muscles more efficiently in order to improve the overall functioning of the body. Examples of the Alexander technique as CAM are using it to treat low-back pain and the symptoms of Parkinson’s disease.

*Feldenkrais method*

The 2012 field representative’s manual defines the Feldenkrais method as a method of education in physical coordination and movement. Practitioners use verbal guidance and light touch to teach the method through one-on-one lessons and group classes. The intent is to help the person become more aware of how the body moves through space and to improve physical functioning.

*Pilates*

The 2012 field representative’s manual defines Pilates as a method of physical exercise used to strengthen and build control of muscles, especially those used for posture. Awareness of breathing and precise control of movements are integral components of Pilates. Special equipment, if available, is often used.

*Trager Psychophysical Integration*

The 2012 field representative’s manual defines Trager Psychophysical Integration as a therapy in which practitioners apply a series of gentle, rhythmic rocking movements to the joints. They also teach physical and mental self-care exercises to reinforce the proper movement of the body. The intent is to release physical tension and increase the body’s range of motion. An example of Trager Psychophysical Integration as CAM is using it to treat chronic headaches.

### Appendix VI – Special diets

*The Atkins diet*

The 2012 field representative’s manual states that The Atkins diet emphasizes a drastic reduction in the daily intake of carbohydrates (40 grams or less), countered by an increase in protein and fat.

*Macrobiotic diet*

The 2012 field representative’s manual states that a macrobiotic diet is low in fat, emphasizes whole grains and vegetables, and restricts the intake of fluids. Of particular importance is the consumption of fresh, non-processed foods.

*The Ornish diet*

The 2012 field representative’s manual states that The Ornish diet is a high fiber, low-fat vegetarian diet that promotes weight loss and health by controlling what one eats, not by restricting the intake of calories. Fruits, beans, grains, and vegetables can be eaten at all meals, while non-fat dairy products such as skim milk, non-fat cheeses, and egg whites are to be consumed in moderation. Products such as oils, avocados, nuts and seeds, and meats of all kind are avoided.

*The Pritikin diet (or Pritikin Principle)*

The 2012 field representative’s manual states that while meat is allowed, the Pritikin diet (or Pritikin Principle) is low-fat and emphasizes the consumption of foods with a large volume of fiber and water, including many vegetables, fruits, beans, and natural, unprocessed grains.

*Vegetarian diet*

The 2012 field representative’s manual defines a vegetarian diet as totally devoid of meat, red or white. There are, however, numerous variations on the non-meat theme. For example, some vegetarian diets are restricted to plant products only, while others may include eggs and dairy products. Another variation limits consumption to raw fruit, sometimes supplemented with nuts and vegetables. Finally, a number of vegetarian diets prohibit alcohol, sugar, caffeine, or processed foods.
